# Computational estimates of mitral annular diameter in systole and diastole cardiac cycle reveal novel genetic determinants of valve function and disease

**DOI:** 10.1101/2020.12.02.20242206

**Authors:** Mengyao Yu, Catherine Tcheandjieu, Adrien Georges, Ke Xiao, Helio Tejeda, Christian Dina, Thierry Le Tourneau, Ina Fiterau, Regeneron Genetics Center, Renae Judy, Noah Tsao, Dulguun Amgalan, Chad J Munger, Jesse M Engreitz, Scott Damrauer, Nabila Bouatia-Naji, James R. Priest

## Abstract

The fibrous annulus of the mitral valve defines the functional orifice and anchors the anterior and posterior leaflets, playing an important role in normal cardiovascular physiology and valvular function. We derived automated estimates of mitral valve annular diameter in the 4-chamber view from 32,220 MRI images from the UK Biobank at ventricular systole and diastole as the basis for genome wide association studies. Mitral annular dimensions correspond to previously described anatomical norms and GWAS inclusive of four population strata identify ten loci, including novel loci (*GOSR2, ERBB4, MCTP2, MCPH1*) and genes related to cardiac contractility (*BAG3, TTN, RBFOX1*). ATAC-seq of primary mitral valve tissue localize multiple variants to regions of open chromatin in biologically relevant cell types and rs17608766 to an algorithmically predicted enhancer element in *GOSR2*. We observed strong genetic correlation with measures of contractility and mitral valve disease, and clinical correlations with heart failure, cerebrovascular disease, and ventricular arrythmias. A polygenic score of mitral valve annular diameter in systole was predictive of risk mitral valve prolapse across four cohorts (Odds ratio 1.19 per SD increase in polygenic score, 95% confidence interval 1.14 to 1.24, p=4.9E-11). In summary genetic and clinical studies of mitral valve annular diameter reveal new genetic determinants of mitral valve biology while highlighting known and previously unrecognized clinical associations. Polygenic determinants of mitral valve annular diameter may represent an independent risk-factor for mitral prolapse. Overall, computationally estimated phenotypes derived at scale from medical imaging represent an important substrate for genetic discovery and clinical risk prediction.

## INTRODUCTION

The mitral valve is a two-leaflet structure anchored to the annulus with secondary chordal attachments to the ventricle which facilitates blood flow from the left atrium to ventricle and prevents regurgitant flow into the left atrium during ventricular systole. The annulus of the mitral valve is an ellipsoidal D-shaped fibrous ring and the anterior-posterior diameter may be measured clinically in the 4-chamber view of cardiac imaging by echocardiogram, computed tomography, or magnetic resonance imaging (MRI)(1).

Both the annulus and leaflets of the mitral valve arise from the atrioventricular cushions during early cardiac development during weeks 5-6 weeks post-fertilization in human cardiovascular development(2, 3).The normal process of valvulogenesis may be disturbed in Mendelian forms of disease or alternately acquired pathology localized to the leaflets or the annulus may compromise mitral valve function resulting in either regurgitation or stenosis(4). In mitral valve prolapse (MVP) one or both leaflets of the mitral valve may become thickened or dysplastic and deviate into the left atrium during ventricular systole typically causing regurgitation, and risk for MVP is influenced by the size of the mitral annulus(5). Recent work has implicated the role of common variation impacting cellular alignment, ciliary function, and TGF-beta signaling in mitral valve prolapse (MVP)(6, 7).

Here we report automated extraction of mitral valve annular diameter at systole and diastole from cardiac MRI data. Extracted data correspond to previously described anatomical norms and GWAS inclusive of four population substrata identify ten loci, including newly described loci and loci previously implicated in mitral valve biology or cardiac contractility. Polygenic scoring of mitral valve annular diameter is predictive of mitral valve regurgitation across four cohorts.

## METHODS

### Ethics statement

This work employed deidentified data from the UK Biobank. Ethical approval was granted by the NHS National Research Ethics Service (ref: 11/NW/0382).

### UK Biobank cohort and mitral valve measurement from cardiac MRI

Using a previously described set of algorithms based upon an open-source deep learning framework(8), we built a U-Net segmentation model with pre-trained weights from VGG11, trained on 60 hand labeled 4-chamber view images, validated on 20 hand labeled images, and tested on 20 hand labeled images which yielded a validation dice score of 91.2% and a the test dice score of 93.8% which were equivalent to differences observed between expert human annotators(8). Then the segmentation model was applied to 32,219 4-chamber images from the UKBB dataset to generate masks for all 4 chambers including left atrium, left ventricle, right atrium and right ventricle. After the masks were generated from the trained U-Net segmentation model, another second measuring function was applied to measure the mitral valve annulus diameter, tricuspid valve annulus diameter, interventricular septum length, and atrial septum length. In this measuring function, first the base of the atriums and apex of the ventricles are found along the medial axis of the chambers, secondly, the intersecting lines of the atriums and the ventricles were used to locate the annuli of the atrioventricular valves from which, annular diameter could be estimated. Finally, measurements were converted to mm from pixels in units using the metadata in the dicom files.

Left ventricular end-systole and left ventricular end-diastole were obtained by selecting the image frame with the largest and smallest estimates of left ventricular volume or left atrial volume as provided by the segmentation algorithm to select the frames best representing left ventricular end-systole and left-ventricular end-diastole.

To exclude outliers related to imaging error or methodological inaccuracy, automated measurements were plotted relative to body surface area with standard measures of quality control. We excluded measures +/-3 standard deviations for each of the four measures. A trained clinician (JRP) performed manual annotation of 50 randomly selected images spanning the cardiac cycle to gauge the accuracy of the estimate of mitral valve annulus diameter.

Manual annotation was performed blinded to automated measures or anthropometric characteristics. The percent difference between automated and manual measurements ([automated measure - manual measure]/manual measure) was examined for systematic relationships to anthropomorphic predictors (Age in years, body surface area, genetic sex), imaging acquisition error (posterior or anterior deviation of the imaging plane from the ideal), and cardiac cycle (ventricular systole or ventricular diastole) using standard linear modeling.

### GWAS and annotation

The UK Biobank data release available at the time of analysis included genotypes for 488,377 participants, obtained through either the custom UK Biobank Axiom array or the Affymetrix Axiom Array. Genotypes imputed to the TOPMed panel (version 5) the Michigan imputation server. Only variants with minor allele frequency (MAF) greater than or equal to 0.01, and minor allele count (MAC) greater or equal to 5, and variants which have Hardy-Weinberg equilibrium exact test p-value upper 1e-20 in the entire MRI dataset and an empirical-theoretical variance ratio (MaCH’s r^2^) threshold above 0.3 were included. The main GWAS was conducted on the largest subset of participants with MRI data from the largest unrelated European-ancestry cohort defined using the variable in.white.British.ancestry.subset in the file ukb_sqc_v2.txt provided as part of the UKB data release (n = 32,220 individuals with estimates derived from imaging data). To replicate the findings, we separated the global dataset into a discovery set and a replication set, which included at least 22,124 and 4,950 participants, separately. To further replicate the findings from the 1st stage, we included three independent sets of other ethnic background with cardiac MRI who were not included in the discovery set, including African/Afro-Carribean (AF, n = 222, age = 49.6±7.0), East Asian (EAS, n = 85, age = 49.2±5.5), and South Asian (SAS, n = 368, age = 52.1±7.9). Examination of those samples according the genetic principal components showed that many were mostly of European ancestry and were unrelated [Figure S1]. Association tests were performed using linear regression PLINK2 (v2.00a2LM) additive model(9, 10), including gender age, adjusted body surface area, and Principal components 1-10 as covariates. Locuszoom was used to generate regional association plots.

Variants were annotated using FUMA, which required liftover conversion of variant coordinates from GRCh38/hg38 to GRCh37/hg19 as input and based on the 1000G Phase3 EUR reference panel population. The independent significant SNPs were identified by the LD r2 0.6. All SNPs in LD with any of independent significant SNPs with LD r2 greater or equal to 0.6 were annotated to eQTL (GTEx v8: heart atrial appendage and heart left ventricle), CADD, RDB, and the GWAS catalog. Linkage disequilibrium (LD) between pairs of variants was examined and reported in R^2^ using the web-based LDPair tool (Release 04/29/2020 https://ldlink.nci.nih.gov).

We analyzed open chromatin regions from mitral valves of 5 patients undergoing mitral valve replacement as previously described(11). Variants in high LD (r2≥0.5 in European populations) with lead SNPs were retrieved using LDproxy function of LDlinkR package(12). Variants overlapping open chromatin in at least two mitral valve samples were reported. We tested overlap of variants with ATAC-Seq peaks (narrowpeak from MACS2 output + 100bp on each side) using bedtools (v2.29.0) annotate function. Variants overlapping open chromatin in at least two mitral valve samples were reported. Read density profiles from two valve datasets, two fibroblasts cell lines and two heart datasets from ENCODE were calculated as previously described(11)) and visualized using Integrated Genome Browser (v9.1.4). We also visualized H3K27ac histone mark read density profiles of ENCODE datasets generated from heart left ventricle (ENCSR702OVJ), ascending aorta (ENCSR982QIF) or primary fibroblasts (ENCSR000APN).

For an analysis of predicted enhancer mapping, we considered the set of variants in linkage disequilibrium (r2 >= 0.8) with the lead variants in 1000 Genomes, and overlapped these variants with predicted enhancers in 131 cell types identified by the ABC Model, which combines measurements of enhancer activity (based on ATAC-seq, DNase-seq, and H3K27ac ChIP-seq) with measurements of 3D contact frequencies (based on Hi-C)(13, 14). We examined DNase-seq and ATAC-seq data across a range of cardiovascular cell types from the ENCODE Project(15). Finally, we examined sequence motif predictions for identified variants using a database of transcription factor binding site motifs(16).

### LDSC and genetic correlation analyses

To calculate genetic correlation between polygenic risk score of mitral valve annular size and other related phenotypes, we obtained summary statistics for cardiac MRI-derived LV measurements (left ventricular end-diastolic volume (LVEDV), left ventricular end-systolic volume (LVESV), stroke volume (SV), the body-surface-area (BSA) indexed versions for cardiovascular traits (LVEDVi, LVESVi, and SVi), and left ventricular ejection fraction (LVEF)(17), atrial fibrillation (AF)(18), nonischemic cardiomyopathy (NICM)(19), heart failure(20), heart failure using UK Biobank data(19), hypertension(21), PR Interval(22), Myocardial Infarction (MI) and coronary artery disease (CAD)(23), heart rate(24), and mitral valve prolapse (MVP)(6). Using these data we performed LD Score regression(25) based on the reformatted summary statistics filtered to HapMap3.

### Phenome wide association study

We performed a phenome-wide association studies (PheWAS) to highlight clinical associations with up to 1,240 ICD-10 aggregated clinical phenotypes defined as phecodes(26). Methods are as previously described(27). For PheWAS results reported here, we excluded phenotypes with less than 50 individuals (for continuous traits) or less than 50 cases (for binary-coded traits) were excluded, and we corrected for multiple testing using a Bonferroni adjustment. We performed PheWAS for the mitral annulus measurements amongst individuals with the MRI data (n=32,219), PheWAS for the GWAS significant variants and for the polygenic scores (described below) amongst individuals without the MRI data (n=308,683 participants).

### Polygenic Prediction of Mitral Valve Prolapse (MVP)

Summary statistics from the mitral valve annular diameter GWAS were used to compute a polygenic score on the remaining set of unrelated European-ancestry subjects. We separated the European ancestry replication set further divided into a training (n=4000) and validation subset (n=1000). We trained the best PRS using the summary statistics from the discovery set in the training subset using PRSice-2(28) and validated the maximally performing PRS (measured by R2) using the validation set. Only variants with MAF greater 0.01 were used to calculate the PRS, as well as the variants in LD r2 0.8. We also validated the best PRS in the MVP cohorts from France, Nantes, UK Biobank, and Penn Medicine Biobank, which include 1007 cases and 1469 controls, 479 cases and 862 controls, 434 cases and 4527 controls, and 144 cases and 20890 controls, respectively. The definition of Mitral valve prolapse in the Penn Medicine Biobank was determined using ICD10 code I34.1 (non-rheumatic mitral valve prolapse), while excluding individuals with codes I05.1, I05.2, 394.1 which incorporate or do not specifically exclude mitral valve dysfunction related to rheumatic heart disease. The association between MVP and the best PRS of mitral valve annular diameter at the MVP individual level were conducted using logistic regression. The sum of best PRS for MVP and MR at the individual level was estimated by PLINK2.

## RESULTS

After exclusion of outliers and individuals +/-2.5 standard deviation, estimates of mitral valve annular diameter were available at two different timepoints during the cardiac cycle (31,973 left ventricular end-systole, 31,864 left-ventricular end-diastole) and conformed to previously described population-based norms(29) [Fig.1A-B]. Automated measures were on average 0.8 mm or 2.2% larger than manual measures with no systematic relationship of the error in mitral valve annular measurement to body surface area, genetic sex, age, or imaging acquisition error observed across 50 randomly selected test images. We performed the main GWAS on mitral valve annular diameter measured at LV systole (n=27,156) and LV diastole (n=27,074) followed by the replication GWAS, where we observed genetic signals meeting standard levels of threshold for genome-wide significance for both Mitral valve annular diameter measured in diastole and systole [Fig.1C-D]. Results from systolic measurements centered around known loci related to cardiac contractility (*TTN*: chr2, rs80182096, p_global_ = 5.37×10^−10^ and *BAG3*: chr10, rs17099139, p_global_= 8.02×10^−09^) as well as novel loci (*ERBB4*: chr2, rs4673661, p_global_= 1.28×10^−10^and an intergenic locus at 6p22.3 proximal to *HDGFL1*: chr6, rs10485012, p_global_ = 6.33×10^−10^) while a set of intronic variants in *GOSR2* at chr17 displayed different lead variants identified in systole (rs6504673, p_global_ = 1.40×10^−12^) and diastole (rs533030436, p_global_= 3.63×10^−13^) which are unlinked (R^2^ 0.02) within European populations [Table 1, Figure S2].

**Figure 1.**
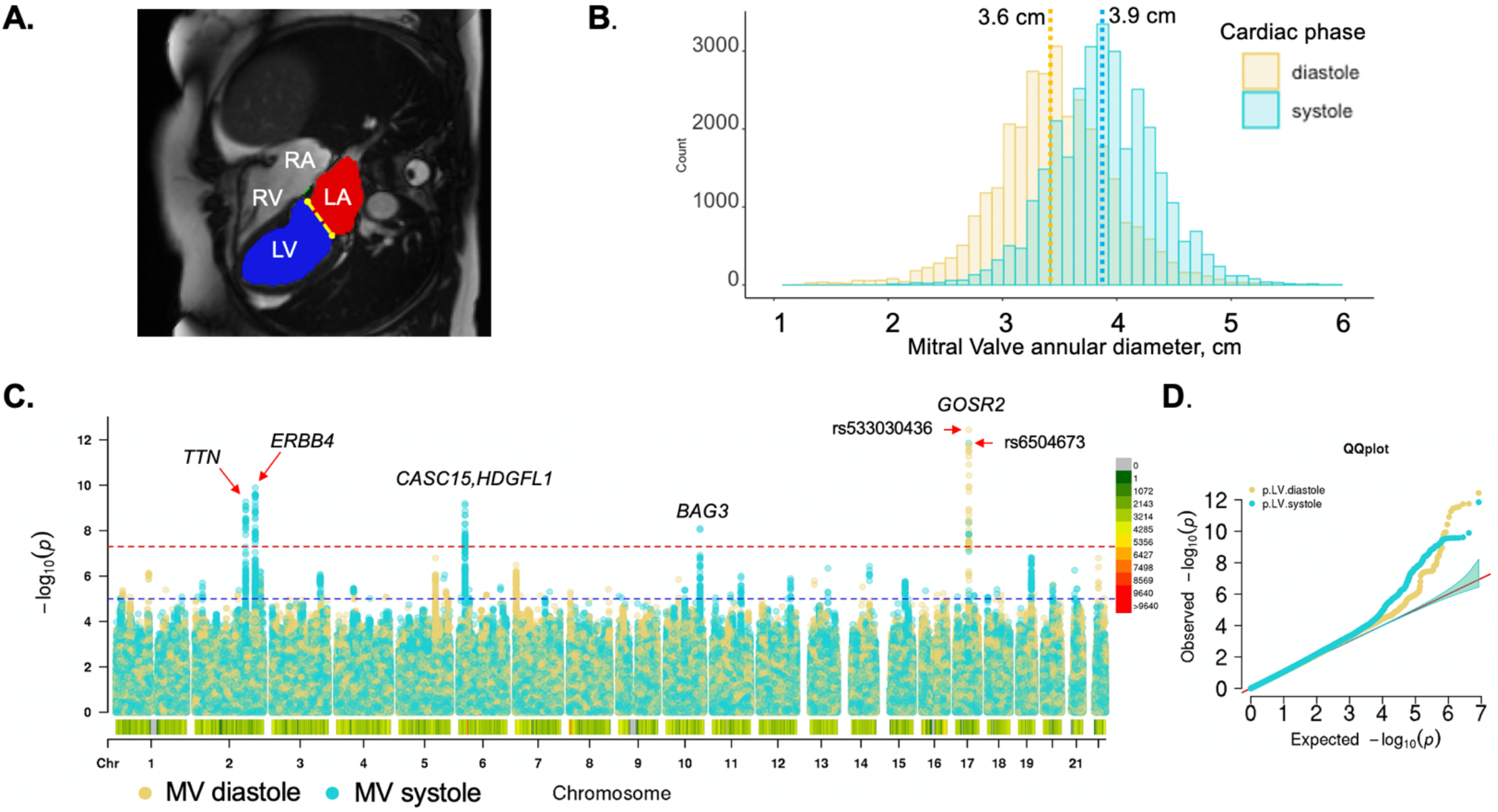
Genome wide association of automated measures of mitral valve annulus diameter. A. Still frame of a cardiac MRI in the four-chamber view demonstrating segmentation of the left ventricle (LV) indicated by the blue region, left atrium (LA) indicated by the red region, and the diameter of the annulus of the mitral valve in the anterior-posterior view (yellow dotted line). B. Population distribution of mitral valve annular diameter estimates in ventricular systole and diastole which conform to previous population-based estimates derived from cardiac MRI. As expected, systolic measurements are greater than diastolic measurements as the annulus experiences deformation during ventricular contraction. Mean values are indicated by dotted lines and labeled. C. Manhattan plot of European-ancestry individuals highlighting unique and shared loci for MV annulus diameter in systole (light blue) and diastole (yellow) D. Quantile-quantile plot indicates an absence of systematic inflation of genetic determinants for both measurements.

**Table 1.**
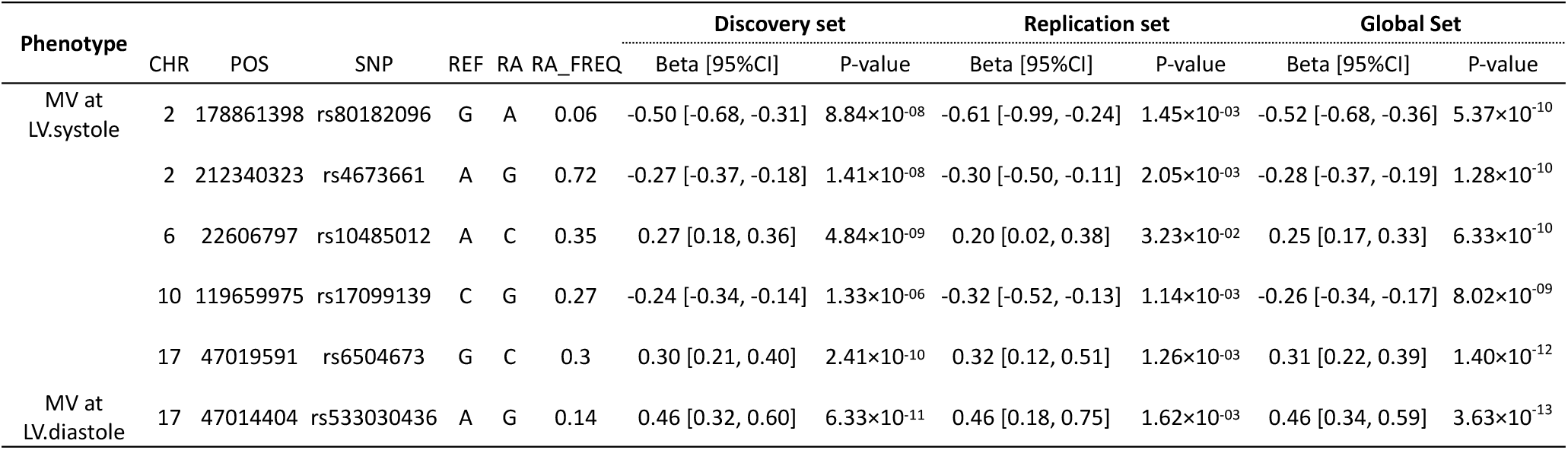
Genome-wide significant results from measurements of mitral valve annulus size at systole and diastole in European ancestry individuals. RA: risk allele, FREQ: frequency

We examined variants overlapping open chromatin regions from mitral valve tissue and a variety of other cell types among replicated lead variants and variants in high LD (r2≥0.5) in European populations. We found overlapping variants at 3 of the 6 loci [Table S1]. At the chromosome 17 locus rs1768766 is in high LD with MV diastolic diameter associated variant (rs533030436) and overlaps open chromatin in primary mitral valve tissue and fibroblasts [Fig.2A] and disrupts a canonical KLF4 binding motif within an activity-by-contact predicted endothelial cell enhancer interacting with the promoter of *GOSR2* [Fig.2B]. At the same locus, an insertion variant from a different haplotype [Figure S9], rs71365052 is in linkage with MV systolic diameter associated variant rs6504673 and overlaps a prominent open chromatin region in mitral valve and fibroblasts, as well as strong active enhancer marks in heart, fibroblasts and aorta [Fig.2C]. Interestingly, both variants appear to be eQTLs of GOSR2 transcripts in fibroblasts (Table S1, Fig.2D), even though rs71365052 is located more than 200kb downstream of the *GOSR2* locus. At chromosome 2 locus proximal to *CCDC141*, one variant (rs60105920), in strong LD with lead SNP rs10485012, overlapped a prominent open chromatin region in valve tissue and fibroblasts, as well as active enhancer histone marks in heart tissue and aorta (Fig.S3A). This variant is an eQTL for *FKBP7* in primary fibroblasts, located more than 300kb upstream of the top associated variants (Fig.S3B).

**Figure 2.**
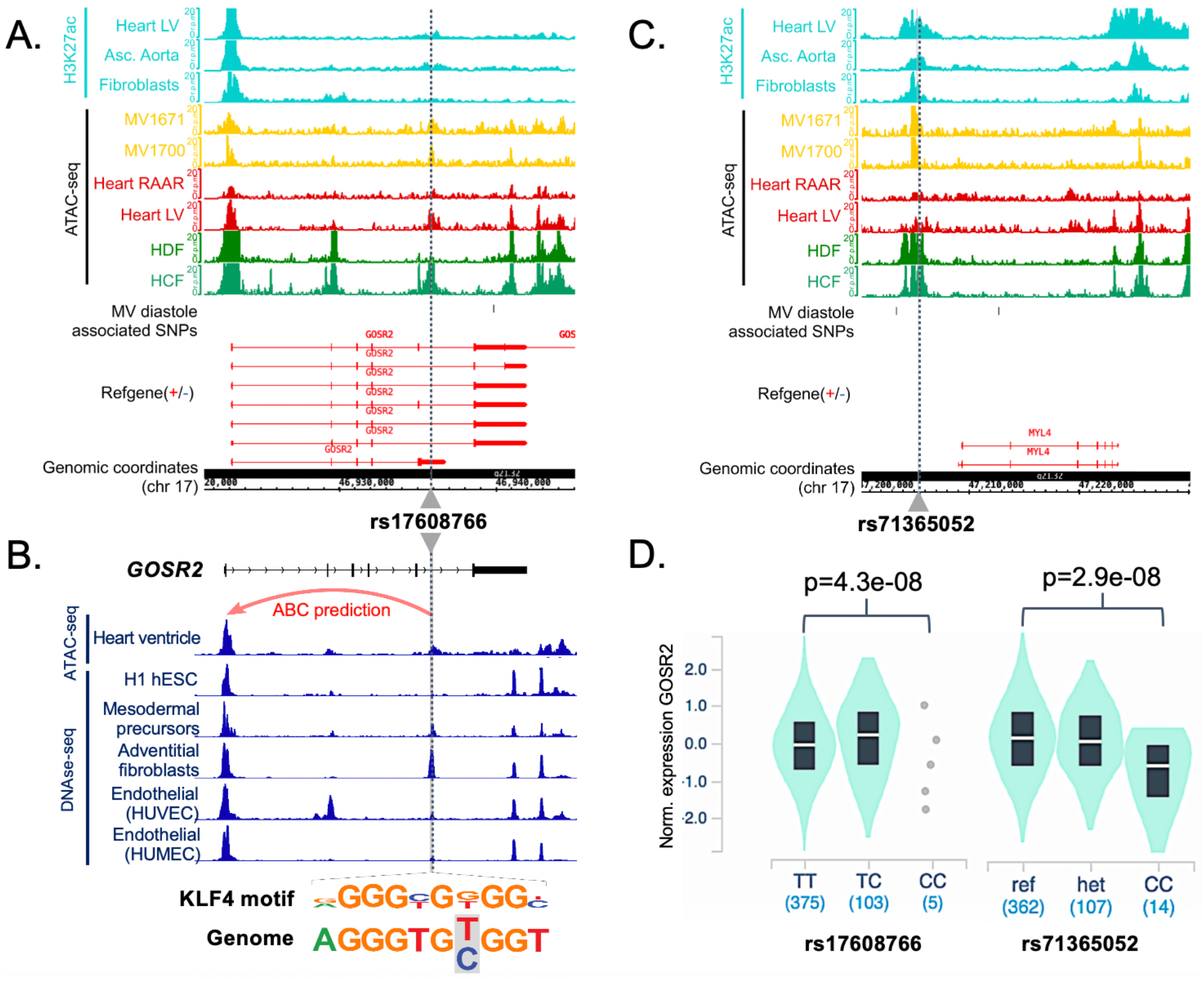
Variation associated with mitral valve annular diameter in systole and diastole is centered in open chromatin regions around *GOSR2*. **A**. Visualization of ATAC-seq/Histone ChIP read densities (in reads/million, r.p.m.) at chromosome 17 locus in the regions surrounding rs17608766 which is associated with mitral valve annular diameter in diastole. Notably the region of open chromatin is present in ATAC-seq data derived from primary mitral valve tissue (MV1671 and MV1700) **B**. The same variant rs17608766 overlaps open chromatin regions present in cardiovascular precursors and endothelial cells but not embryonic stem cells (hESC) and occurs within an activity-by-contract (ABC) enhancer element predicted to interact with the promoter of *GOSR2* in heart ventricle tissue and endothelial cells (red arrow) and specifically disrupts a KLF4 binding motif. **C**. ATAC-seq/Histone ChIP read densities for the locus of rs71365052 associated with mitral valve annular diameter in systole which appears in a large region of open chromatin proximal to *MYL4*. **D**. Violin plot representation of association between genotype and GOSR2 expression in cultured fibroblasts for rs17608766 and rs71365052 derived from GTEx (v8 release) shows strong relationship with GOSR2 for both variants in opposite directions. T (r2 = 0.03). Both risk alleles (rs1768766-C and rs71365052-ref) are associated with higher expression of GOSR2 and larger mitral annular diameter. Abbreviations MV: Normal Valves. HDF: human dermal fibroblasts. HCF: human cardiac fibroblasts. Heart LV: Heart left ventricle. Heart RAAR: Heart right atrium auricular region. Asc.Aorta: Ascending Aorta. HMVEC: human microvascular endothelial cells. HUVEC: human umbilical endothelial cells.

We also performed a trans-ethnic meta-analysis [Figure S3], which revealed new loci meeting genome-wide significance for mitral valve annulus diameter at diastole (*MCTP2, MCPH1*) and systole (*MROH7*, and an intergenic locus at 16q21) with lead variants discovered amongst the African/Afro-Caribbean population strata [Figures S4 & S5]. Candidate signals of interest were observed in mitral valve measurements from the two smaller population strata of South and East Asian descent but did not meet standard significance thresholds in the trans-ethnic meta-analysis [Table S1].

Genome-wide significant variants for systole were each nominally significant (p < 0.05) for diastole and vice versa [Table S2], and LD-score correlation (LDSC) confirmed a substantial genetic overlap in the genetic basis of the two measurements of mitral annulus diameter as would be expected in measures derived from the same tissue in the same set of individuals (p=8.15E-83) [Figure 3]. In addition, for measures of mitral annulus diameter at systole and diastole, LDSC suggested a strong positive genetic correlation with GWAS of left ventricular volumes, stroke volume, and ejection fraction derived from analyses of a largely overlapping dataset(17) and a strong negative correlation with ejection fraction, heart rate [Figure 3]. Of note, mitral valve prolapse (MVP) displayed a negative genetic correlation with mitral valve annulus measurement at systole (p = 7.42e-06) while atrial fibrillation displayed a positive genetic correlation with annulus measurement at diastole (p = 1.66e-06).

**Figure 3.**
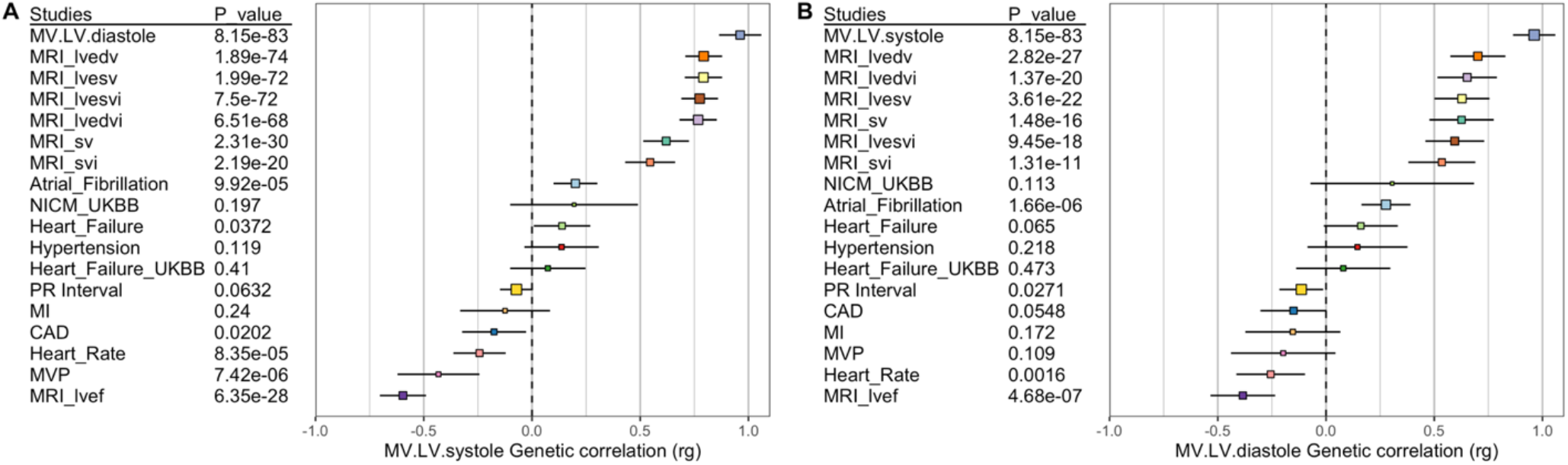
Forest plots showing genetic correlation of mitral valve annular diameter with related cardiovascular phenotypes. A. For annular diameter measured at systole there is a strong positive correlation with indexed measures of left ventricular volume obtained from a largely overlapping dataset as well as with atrial fibrillation, and a negative correlation with heart rate, mitral valve prolapse and ejection fraction. B. Annular diameter measured at diastole displays many of the same correlations, as well as a positive correlation with atrial fibrillation. Abbreviations: lv-left ventricular, edv-end diastolic volume, esv-end systolic volume, sv-stroke volume, i-indexed to body surface area, ef-ejection fraction, NICM -- non-ischemic cardiomyopathy, MI – myocardial infarction, CAD—coronary artery disease, MVP—mitral valve prolapse.

To better understand the clinical correlates of our anatomical estimates and genetic findings, we performed PheWAS of mitral annular diameter amongst the individuals with a measurement. Amongst the 32,219 individuals with a measurement, a larger annular diameter in diastole and systole were positively associated with heart failure and cardiomyopathy, while smaller annular diameter in both measures were associated with Type 2 diabetes, hyperlipidemia, and hernias affecting the diaphragmatic surface of the heart [Fig.4A]. We also performed a PheWAS of four lead variants identified in the GWAS within individuals without a mitral valve measurement (n = 308,683) which suggested that the C allele of rs10485012 (associated with larger annular diameter in systole) was associated with decreased risk of coronary atherosclerosis and heart failure, while the G allele of rs533030436 (associated with larger annular diameter in diastole) specifically displayed an association with decreased risk for cerebrovascular disease [Fig.4B]. Interestingly, the PheWAS for mitral valve annular diameter in diastole specifically highlighted the direct association with non-rheumatic disorders of the mitral valve [Fig.4A]. Therefore, we created a polygenic score for mitral valve annular diameter for the diastolic and systolic measurements using the validation dataset (described above) subdivided into two groups which captured a larger amount of variation for the systolic measurement (R2 = 0.011, p=6.7E-11) than the diastolic measurement (R2 = 0.006, p = 3.1E-07) [Figure S6]. Across four separate cohorts of mitral valve prolapse the polygenic score from both measures behaved as expected with the mitral annular score from systole captured a larger proportion of R2 than the diastolic measure. The polygenic score for mitral valve annular diameter measured at ventricular systole displayed the strongest prediction of risk for mitral valve prolapse (MVP) or regurgitation ranging from an odds ratio (OR) of 1.14 to 1.31 between the different cohorts [Figure 5]. In meta-analysis across the four cohorts, per standard deviation increase in polygenic score of mitral valve annular diameter at systole there was a 1.1 fold increase in the risk of MVP (OR 1.19, 95% confidence interval (95%CI) 1.14 to 1.24, p=4.9E-11), while the risk related to mitral valve annular diameter at diastole was lower (OR 1.13, 95% CI 1.07 to 1.18, p = 1.38E-05) [Figure 5]. For each standard deviation increase in the polygenic score for mitral annular diameter at systole we observed a small increased risk of mitral valve regurgitation in the Penn Medicine Biobank which did not meet nominal thresholds for statistical significance [Figure S7]. Finally, using the polygenic scores generated we also performed a PheWAS within UK Biobank individuals without imaging data (n = 308,683). The PRS for mitral valve annular diameter at diastole was positively associated with varicose veins and ventricular tachycardia [Figure S8], while inversely associated with Type 2 diabetes.

**Figure 4.**
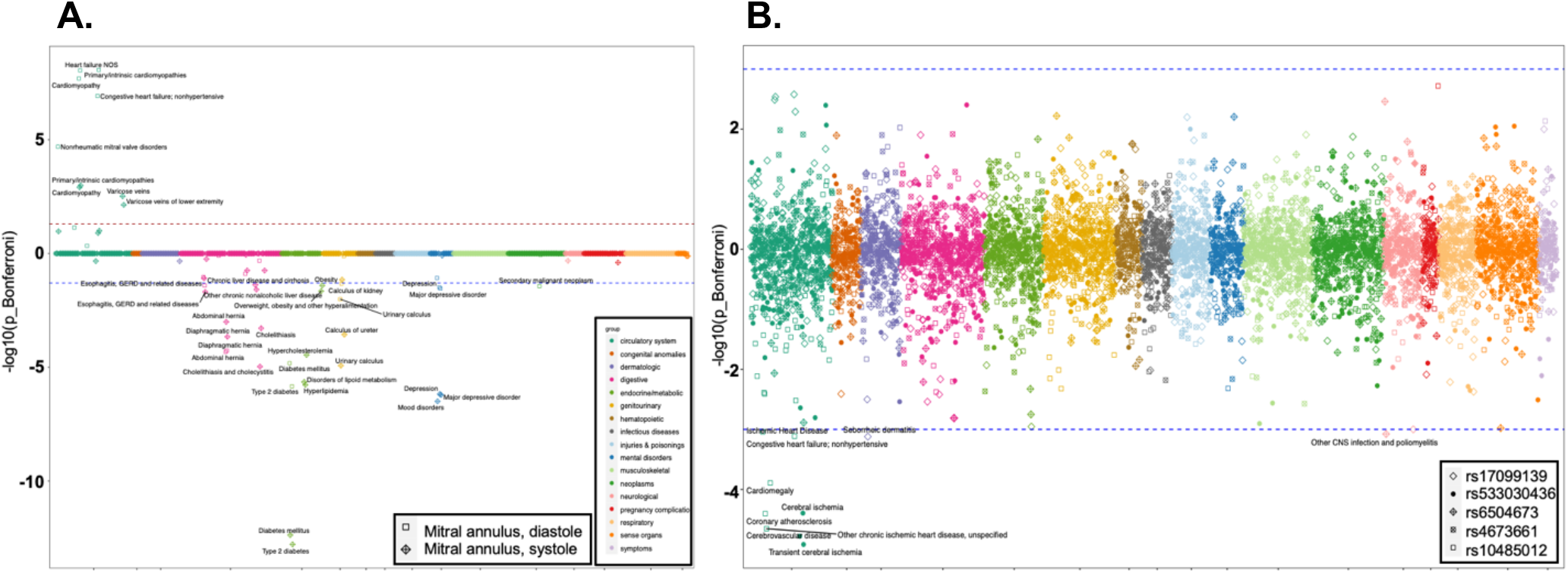
PheWAS of estimated measures of mitral annulus size and variants identified in GWAS identifies clinical correlates of disease. **A**. PheWAS of mitral annular diameter amongst the individuals with a measurement, the absolute value of Y-axis represents the negative logarithm of the p-value with values above and below zero representing positive and negative correlations respectively. Colors represent different clinical/anatomical categories for the phecodes used. Larger annular diameter measured in diastole and systole are positively associated with heart failure and cardiomyopathy, while smaller annular diameter in both measures were associated with Type 2 diabetes, hyperlipidemia, and hernias. **B**. We also performed a PheWAS of five lead variants identified in the GWAS within individuals without a mitral valve measurement (n = 308,683) which suggested that the C allele of rs10485012 (associated with larger annular diameter in systole) was associated with decreased risk of coronary atherosclerosis and heart failure, while the G allele of rs533030436 (associated with larger annular diameter in diastole) specifically displayed an association with decreased risk for cerebrovascular disease phenotypes.

**Figure 5.**
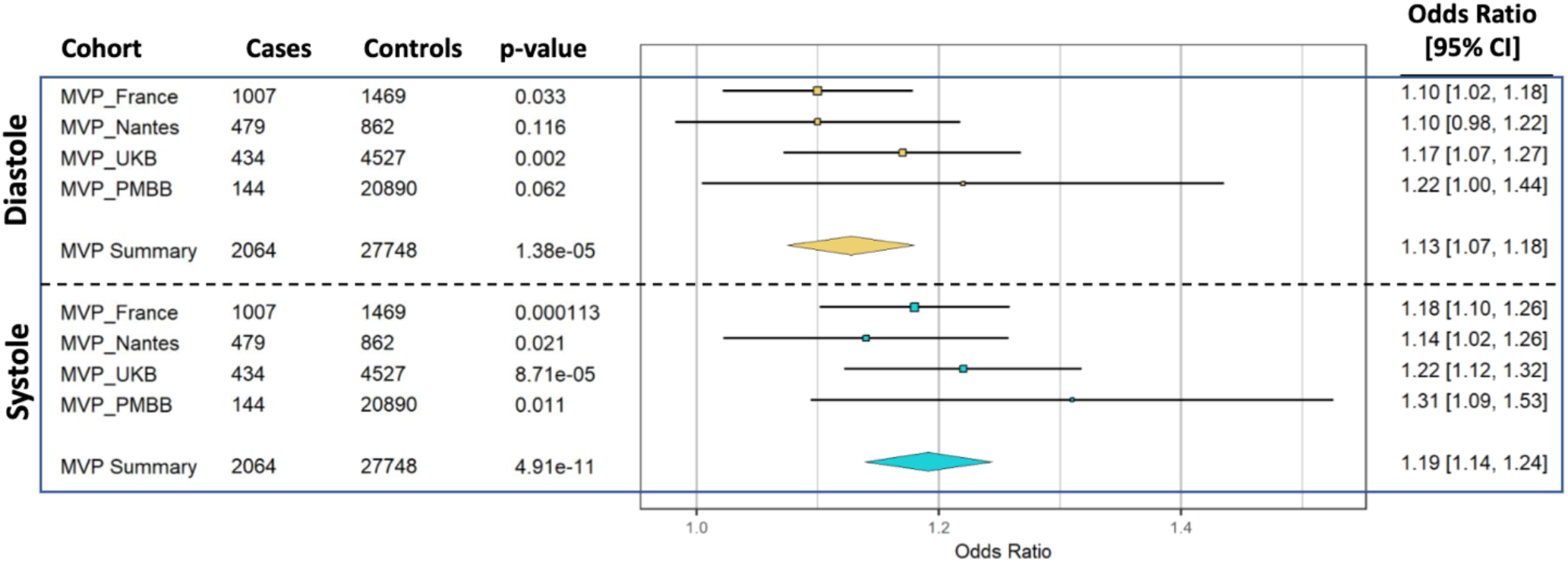
Forest plots relating the polygenic risk score for mitral valve annular diameter measured in systole and diastole to the risk of mitral valve prolapse across four independent cohorts.

## DISCUSSION

Here we have derived automated measures of mitral valve annular diameter at systole and diastole for use in discovery of genetic determinants of mitral valve biology from the UK Biobank. We observe a series of genetic loci which appear to be related directly to mitral valve biology or arising from secondary dilation of the mitral valve annulus related to cardiac contractility. A polygenic score derived from these results is predictive of mitral valve prolapse across multiple cohorts and PheWAS confirms clinical correlates of mitral valve disease. Importantly, our results do not appear to overlap with previous genetic determinants of mitral valve annular calcification(30) [Table S4], suggesting the estimates of mitral annular diameter and genetic results presented here represent an underlying phenotype which does not originate primarily from degenerative or calcific processes associated with aging or turbulent blood flow(31). Importantly, we observed strong genetic signal in the African/Afro-Caribbean population strata which differed from the European population, a finding that underscores the desperate need for genetic studies of cardiovascular phenotypes across the diversity of human populations worldwide.

The majority of loci identified appear to be related directly to development and function of the mitral valve apparatus. The *ERBB4* gene is a receptor tyrosine kinase which responds to epidermal growth factors and is recognized to be essential for normal development of cardiac valves in knockout mice(32). Copy number variation encompassing *MCTP2* has been shown to cause congenital heart disease inclusive of mitral atresia or stenosis, and absent endocardial cushions were observed in morpholino knockdowns of *Mctp2* in *Xenopus* embryos(33). The *HDGFL1* locus has been previously implicated in QRS duration and the PR interval (22, 34) which may be epiphenomenon related to the insulating role of the mitral valve annular tissue in electromechanical coupling of the cardiac action potential(35, 36). Among the other loci highlighted, *MCPH1* has been implicated in blood pressure and carotid intima media thickness, while the intergenic 16q21 locus and *MROH7* appear to be without described direct relationships to cardiac, valvular, or vascular phenotypes.

Previously identified in GWAS of blood pressure(37, 38) and aortic root diameter(38), the two variants in *GOSR2* identified in systole (rs6504673) and diastole (rs533030436) display essentially no linkage in European populations (R2 0.02) and arise from different haplotypes [Figure S9]. The variant rs533030436 identified in diastolic measures is in strong linkage with rs11874 (R^2^ 0.84) recently identified in congenital cardiovascular malformations(39) as well as rs17608766 (R^2^ 0.76) identified in Aortic valve stenosis(27) in the same UK Biobank MRI dataset. The cusps and annulus of the mitral valve are composed primarily of endothelial cells and valvular interstitial cells which may differentiate to myofibroblasts in disease states(40). Our analysis of open chromatin in related cell types (endothelial, cardiomyocyte, fibroblast) is aligned with open chromatin in primary mitral valve tissue. When combined with tissue-specific predictors of 3-dimensional genome confirmation for one example, variants appear to be acting to regulate *GOSR2*. Including the findings presented here, genetic variation in *GOSR2* is now specifically implicated in multiple studies of aortic and mitral valve biology and congenital heart disease which, when taken together with chromatin accessibility data presented here, strongly suggest a primary role in the formation, growth and functional maintenance of cardiac valves and vascular tissue.

While the mitral annulus changes in size during the cardiac cycle, it does not have intrinsic contractile properties(41). Some of the loci identified have are directly related to cardiac contractility. A missense variant in *BAG3* variant rs2234962 resulting in a Cysteine to Arginine substitution at position 151 (CADD score = 21) is linked with the lead variant rs17099139 (R2 =0.74). Both *BAG3* and *TTN* contribute to the function of the contractile apparatus in cardiomyocytes and rare deleterious variation cause Mendelian forms of dilated cardiomyopathy. Common variation in these genes has been recently recognized to directly influence contractility phenotypes within the UK Biobank cardiac MRI data(17). Additionally *RBFOX1* has a well-described role in regulating alternative splicing within cardiomyocytes to mediate contractile changes in heart failure and thus is likely playing an incidental role in mitral valve biology(42). The influence of these variants upon mitral valve annulus size is likely secondary to their impact upon ventricular contractility; dilation of the mitral valve annulus is a direct consequence of dilation of the left ventricle due to decreased systolic performance.

The polygenic score derived from GWAS of mitral annulus size at systole was associated with mitral valve regurgitation or prolapse across four populations, suggesting that the genetic signals derived from anatomical measurements are relevant for the practical purposes of understanding clinical disease. A small amount of mitral valve dysfunction is a clinically insignificant finding or may commonly arise as a consequence of ischemic cardiomyopathy(43, 44). We note that the polygenic risk score for annular diameter was specifically predictive of mitral valve prolapse [Figure 4] and not with mitral regurgitation [Figure S7]. The use of polygenic scores requires extensive validation, before being used clinically for screening and identification of patients at higher risk for adverse remodeling of the left atrium and ventricle which occurs with progression to significant mitral valve dysfunction. Furthermore, the clinical application of such scores is limited by the notable deficiency of genetic data combined with measurements of cardiac imaging in diverse populations(45).

The clinical associations revealed by PheWAS of mitral annular measures, lead GWAS variants, and polygenic scores are broadly consistent with the genetic correlation results and a current clinical understanding of mitral valve physiology. Decreasing cardiac function may cause dilation of the annulus and mitral valve dysfunction, while mitral valve dysfunction whether primary or secondary due to rheumatic fever or papillary muscle rupture may often result in decreased cardiac function. Interestingly, in both the PheWAS of mitral valve measurements and the polygenic scores, psoriasis displayed a negative association with mitral annular diameter in systole, and mitral valve dysfunction has been reported in patients affected with psoriasis(46).

The observational association between mitral valve prolapse and ventricular arrhythmias(47) is supported by the positive association between the PRS for mitral valve annulus diameter in diastole with paroxysmal ventricular tachycardia [Fig.S8], which may suggest that the mitral valve annulus links these two phenotypes. However, we wish to state clearly that while PheWAS results provide context for our findings they do not establish a causal link between mitral valve annular dimensions and the clinical phenotypes identified.

Our study is not without limitations. While we took steps to estimate the error related to image acquisition and our method of automated estimation and to exclude outliers, at each step systematic errors may be introduced which may bias the measurements. Our approach to measuring the mitral annulus is a simple measure derived from a two-dimensional image, other approaches to quantifying mitral valve morphology may provide a better anatomical substrate for interrogation of genetic and polygenic predictors of disease(48). While the anatomical measure of mitral valve annulus size is predictive of valvular function, GWAS on direct measures of valve function in larger numbers of patients from diverse genetic backgrounds are likely to yield additional important insights into the genetic basis of mitral valve biology.

In conclusion, the genetic studies of Mitral valve annular diameter presented here reveal new determinants of mitral valve biology and highlight shared underlying biology with cardiac contractility. Clinical studies highlight both known and previously unrecognized phenotypic associations, and our analyses suggest that polygenic prediction of mitral valve annular diameter may represent an independent risk-factor for mitral prolapse. Importantly our findings highlight the need for inclusion of diverse populations in genetic studies of cardiovascular disease while demonstrating the powerful paradigm of imaging-derived phenotypes for the genetic and clinical studies in cardiovascular health and disease.

## Data Availability

UK Biobank data is available to the public. Penn Biobank and French data from GWAS of Mitral Prolapse are available from the primary authors of those studies upon request.

**Supplementary Figures and Tables for Yu et al**.

(In order of appearance in the main manuscript text)

